# Pre-exposure prophylaxis uptake among users of non-occupational post-exposure prophylaxis: a longitudinal analysis of attendees at a large sexual health clinic in Montréal

**DOI:** 10.1101/2020.04.22.20075614

**Authors:** Yiqing Xia, R. Zoë Greenwald, M. Rachael Milwid, Claire Trottier, Michel Boissonnault, Neil Gaul, Louise Charest, Gabrielle Landry, N. Navid Zahedi, Jason Szabo, Réjean Thomas, Mathieu Maheu-Giroux

## Abstract

**Background:** Reducing HIV transmission using pre-exposure prophylaxis (PrEP) requires targeting individuals at high acquisition risk, such as men who have sex with men (MSM) with a history of non-occupational post-exposure prophylaxis (nPEP). This study aims to characterize longitudinal trends in PrEP uptake and its determinants among nPEP users in Montréal.

**Methods:** Eligible attendees at *Clinique médicale l’Actuel* were recruited prospectively starting in October 2000 (nPEP) and January 2013 (PrEP). Linking these cohorts, we characterized the PEP-to-PrEP cascade, examined the determinants of PrEP uptake after nPEP consultation using a Cox proportional-hazard model, and assessed whether PrEP persistence differed by nPEP history using Kaplan-Meier curves.

**Results:** As of August 2019, 31% of 2,682 MSM nPEP cohort participants had two or more nPEP consultations. Subsequent PrEP consultations occurred among 36% of nPEP users, of which 17% sought nPEP again afterwards. Among 2,718 PrEP cohort participants, 46% reported previous nPEP use. Among nPEP users, those aged 25-49 years (Hazard Ratio (HR)=1.3, 95% confidence interval (CI): 1.1-1.7), with more nPEP episodes (HR=1.4, 95%CI: 1.3-1.5), reported chemsex (HR=1.3, 95%CI: 1.1-1.7), with a STI history (HR=1.5; 95%CI: 1.3-1.7), and who returned for their first nPEP follow-up visit (HR=3.4, 95%CI: 2.7-4.2) had higher rates of PrEP linkage. There was no difference in PrEP persistence between PEP-to-PrEP and PrEP only participants.

**Conclusion:** Over one-third of nPEP users were subsequently prescribed PrEP. However, the large proportion of men who repeatedly use nPEP calls for more efficient PrEP-linkage services and, among those that use PrEP, improved persistence should be encouraged.

## Introduction

In 2016, there were an estimated 2,165 new HIV infections in Canada, of which approximately half occurred among gay, bisexual, and other men who have sex with men (gbMSM) [1]. Worldwide, the risk of HIV infection is 27 times higher in gbMSM than the general population [2]. Addressing the unmet prevention needs of key populations at high risk of HIV acquisition and transmission -such as gbMSM-remains a priority. In 2017 Montréal signed the Paris declaration and, shortly after, became the first Canadian city to adopt the UNAIDS Fast-Track City target of zero new HIV infections by 2030 [3]. This ambitious objective will require strengthening the HIV treatment and care cascade and, crucially, must address unmet HIV prevention needs.

Non-occupational post-exposure prophylaxis (nPEP) and pre-exposure prophylaxis (PrEP) are two strategies for reducing the risk of HIV acquisition which, when combined with other intervention strategies, could have the potential to achieve HIV elimination [4]. nPEP involves taking a 28-day course of highly active antiretroviral tri-therapy following an event that could carry a moderate/high risk of acquisition (initiated within 72 hours of exposure). It was first implemented following occupational exposure in the early 1990s and was later extended to non-occupational situations such as sexual contacts [5]. In contrast, PrEP was recommended by the World Health Organization in 2012 as a strategy for adults who are at a high, ongoing risk of HIV infection [6]. It can be taken daily, or for gbMSM, intermittently (known as “*on demand PrEP*”) [4 7 8]. Randomized control trials among gbMSM have estimated an overall PrEP effectiveness of 86% [7 9-11] for preventing sexual transmission which could reach 99% for those who are perfectly adherent to treatment guidelines [12].

Contrary to the use of nPEP as an emergency measure, it is recommended that those who seek nPEP repeatedly, or are at an ongoing risk of HIV acquisition, be evaluated for possible PrEP use [13-16]. However, emerging evidence suggests that uptake of PrEP among gbMSM who have repeatedly used nPEP is suboptimal [17]. With the Fast-Track city goal of zero new HIV infections, it is crucial to limit these missed prevention opportunities. In 2013, Québec became the first Canadian province to recommend PrEP and fund it using public budgets. Using 6 years of longitudinal data on sexual health clinic attendees, the overarching aim of this study is to characterize the PEP-to-PrEP corridor and identify potential barriers to linkage. Specifically, we will: 1) describe longitudinal trends in nPEP and PrEP use, 2) estimate the time from nPEP to PrEP and the determinants of PrEP uptake by nPEP users, and 3) compare the differences in PrEP persistence between participants with and without a history of nPEP use. These results could help limit missed HIV prevention opportunities among gbMSM.

## Methods

### Study setting

*Clinique médicale l’Actuel* (l’Actuel) is a Montréal-based, sexual health clinic specializing in HIV treatment and prevention, predominantly among gbMSM. The clinic is one of the largest nPEP and PrEP providers in Canada and was one of the first clinics to prescribe nPEP (August 2000) and PrEP (January 2013). Attendees at l’Actuel seeking nPEP and PrEP were recruited prospectively from August 2000 for the nPEP cohort and January 2013 for the PrEP cohort. At each nPEP and PrEP consultation, patients complete a questionnaire regarding their demographic and behavioural characteristics and a consent form. The clinical component of the survey is administered by a nurse or physician. Participants consulting for nPEP were scheduled for follow-up visits with a clinician at 4 and 12 weeks following their initial nPEP prescription, which included HIV testing to monitor nPEP efficacy as well as sexual health and HIV prevention counselling, including PrEP referrals when appropriate. All participants consulting for PrEP were scheduled for a follow-up visit 30 days after the baseline consultation and at 3-month intervals thereafter. De-identified questionnaires were then entered into prospective nPEP and PrEP cohort databases. A detailed description of the cohorts and clinical protocols can be found elsewhere [18 19].

### Study population

The study population included self-identified gbMSM who consulted for nPEP and/or PrEP at *Clinique médicale l’Actuel*. Participants who were HIV positive at their baseline visit were excluded from the study population. The first analysis assessed longitudinal trends in nPEP and PrEP consultations from October 2000 – August 2019.

We restricted the analysis to the time periods during which PrEP was recommended (January 1^st^, 2013 and August 31^st^, 2019) for the primary analysis measuring the determinants of PrEP uptake among nPEP users. Participants were categorized into 4 groups: 1) those who consulted for or received nPEP only (hereafter referred to *as PEP only participants*), 2) those who consulted for or received PrEP only (hereafter referred to *as PrEP only participants*), 3) participants who consulted for or received nPEP prior to using PrEP at l’Actuel (hereafter referred to *PEP-to-PrEP participants*), and 4) PEP-naïve participants who initiated PrEP and subsequently consulted for nPEP after PrEP discontinuation (hereafter referred to *PrEP-to-PEP participants*) (**Figure 1**). Our study design enables us to link participants in the nPEP and PrEP cohorts but not those who sought care at other clinics. As such, we could not properly estimate time to linkage, and the small number of participants who sought nPEP at l’Actuel but PrEP elsewhere (N=13) and PrEP at l’Actuel but nPEP from another clinic (N=120) were excluded from the analyses. Participants whose last nPEP consultation took place before 2013 were also excluded from the study regardless of whether they consulted for PrEP as PrEP was unavailable at that time.

**Figure 1.**
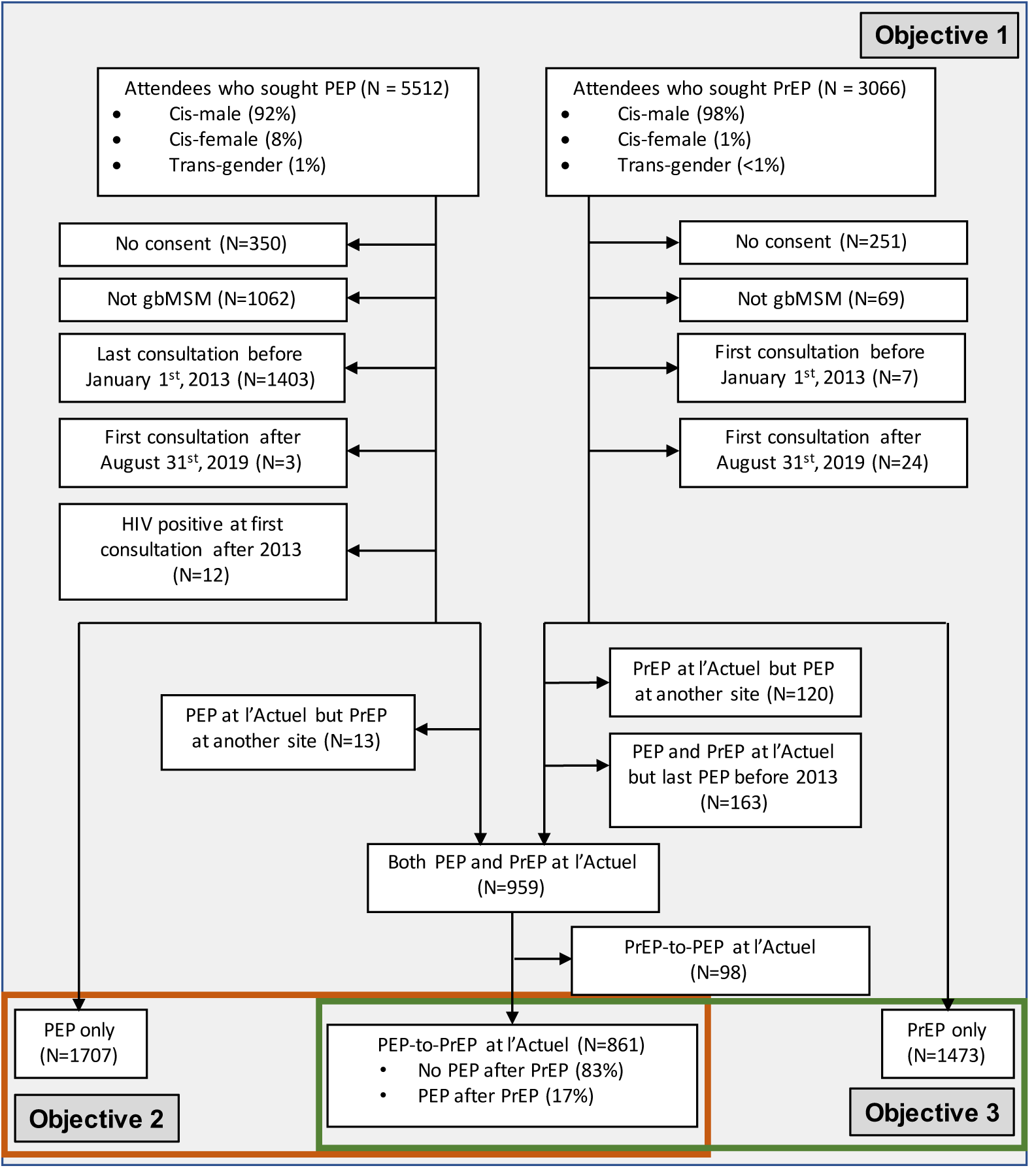
Inclusion and classification of study participants among attendees at the *Clinique médicale l’Actuel*, Montréal, Canada (October 2000 (PEP) / January 2013 (PrEP) to August 2019). Gay, bisexual, and other men who have sexwith men (gbMSM) participants who only consulted for or received non-occupational post-exposure prophylaxis (PEP) or pre-exposure prophylaxis (PrEP) were defined as “*PEP only*” and “*PrEP only*”, respectively. Participants who consulted for or received PrEP subsequently to consulting for nPEP were classified as “*PEP-to-PrEP*”. Participants who consulted for or received nPEP after PrEP were defined as “*PrEP-to-PEP”*. Objective 1 includes all participants, Objective 2 (red box) includes *PEP only* and *PEP-to-PrEP* participants, and Objective 3 (green box) includes *PrEP only* and *PEP-to-PrEP* participants.

### Analyses

Longitudinal trends in nPEP and PrEP consultations from 2000-2019 are presented and the demographic and behavioral profiles of *“PEP only*”, “*PrEP only*”, and “*PEP-to-PrEP*” participants over the 2013-2019 period are compared. The variables examined include: age, education, total number of nPEP episodes, average time between nPEP episodes (for participants who had multiple nPEP episodes and nPEP visits before the first PrEP consultation), adherence to a nPEP treatment (missing <4 pills), early termination of nPEP treatment and whether the participant was re-exposed to HIV during nPEP treatment, chemsex (i.e., sexualized substance use), and a self-reported lifetime history of sexually transmitted infections [17].

The time from the first nPEP visit after 2013 to the first PrEP consultation, stratified by age group (<25, 25-49, ≥50 years, to ensure a large enough sample size in each group while allowing salient age features to be considered), by the total number of nPEP episodes and by the calendar year of the first nPEP visit was explored using Kaplan-Meier curves. Determinants of the time to PrEP uptake among nPEP users were examined using a Cox proportional-hazards model. The model included covariates for which there was previous evidence of potential association [20] or an indication for PrEP [4], including: age and education (secondary and lower, college (including Quebec’s *Collège d’enseignement général et professionnel* (CEGEP) program, university). Additionally, the total number of nPEP episodes, chemsex during the risk episode, self-reported lifetime STI history (any/none) and whether the participants returned for the first nPEP follow-up visit were included in the model and were treated as time-dependent covariates. The year of the first nPEP consult after 2013, was used to stratify the survival analyses. All cohort participants were censored if they seroconverted during follow-up, in August 2019, or at their first PrEP consult, whichever occurred first. Schoenfeld residuals were used to verify that the proportional hazard assumption was met for each predictor-outcome pair [21], while the Efron method was adopted to handle potential ties [22]. Missing values of education (N=325) and STI history (N=91) were handled using multiple imputations and estimates from 5 imputations were pooled using Rubin’s rules [23].

Lastly, a Kaplan-Meier analysis was used to characterise the differences in PrEP persistence between *PEP-to-PrEP* and *PrEP only* participants who initiated PrEP. PrEP termination was defined as a discontinuation of PrEP as indicated by the participant, or as no active PrEP prescription within the past 6 months after the participant’s last PrEP follow-up visit. Participants who consulted for PrEP but who did not attend the first PrEP follow up visit were excluded from the analysis.

All analyses were performed with R version 3.6.1.

Ethics approval was obtained through Veritas Institutional Review Board and McGill University.

## Results

### Description of the study participants

#### Entire l’Actuel cohort

Between October 2000 (for the nPEP cohort) or January 2013 (for the PrEP cohort) and August 2019, 5,512 attendees consulted for nPEP and 3,066 individuals consulted for PrEP at l’Actuel. A total of 6,039 nPEP consultations and 3,253 PrEP consultations took place at l’Actuel among 4,139 and 2,814 gbMSM, respectively. A steady increase in consultations was observed prior to 2016, followed by decreases in both the numbers of nPEP and PrEP consultations from 2017 onwards (**Figure 2**). The simultaneous reduction in nPEP and PrEP can be explained by the emergence of new clinics specializing in HIV that provide similar services.

**Figure 2.**
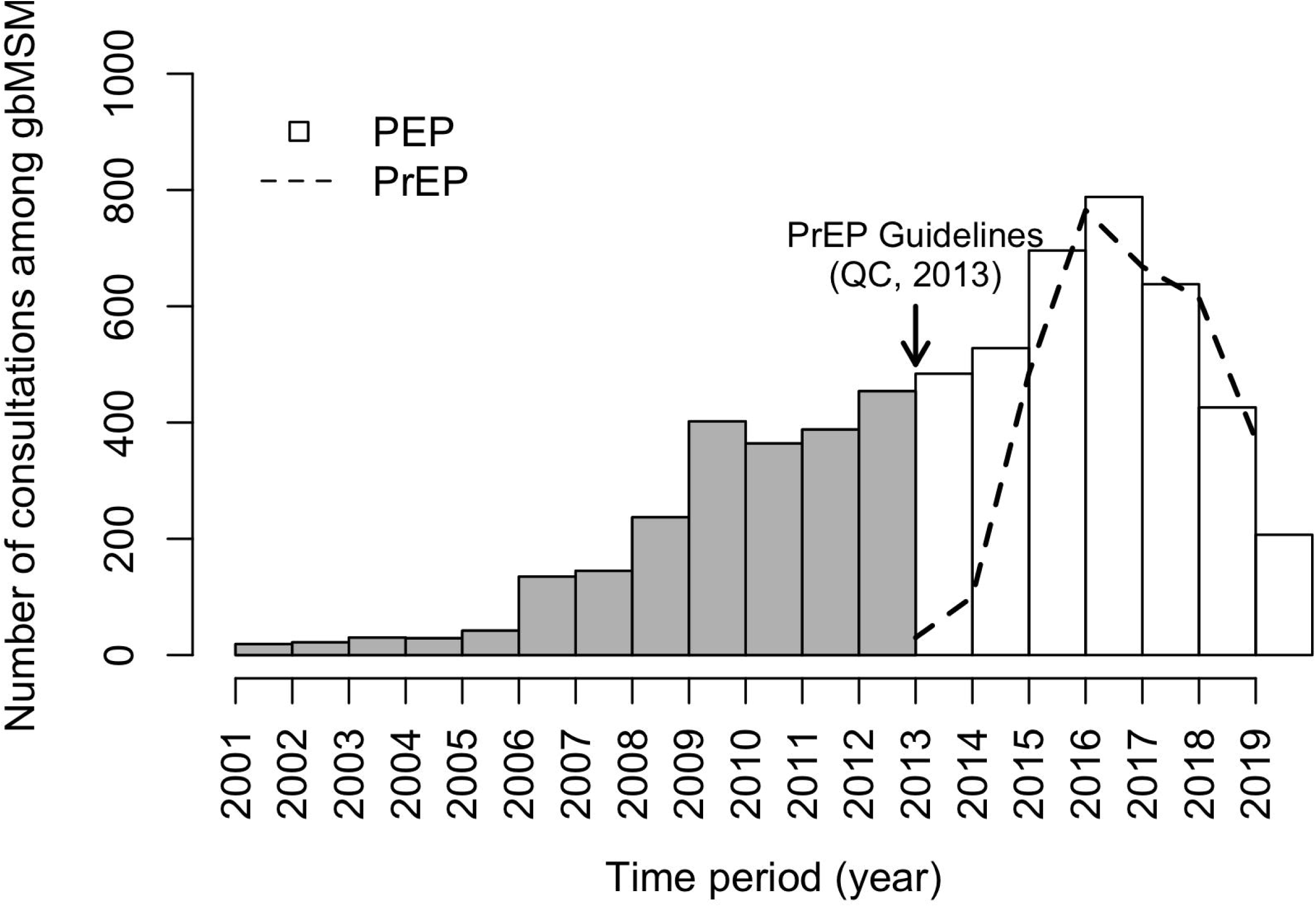
Number of post-exposure prophylaxis (PEP) and pre-exposure prophylaxis (PrEP) consultations conducted among gay, bisexual and other men who have sex with men (gbMSM) between January 2001 and August 2019 at Clinique médicale l’Actuel, Québec (QC), Canada. Each bar represents total number of nPEP consultations during a calendar year with the exception of the last bar which represents an eight-month period. The dashed line represents the total number of PrEP consultations over each time interval between July 2013 and August 2019. nPEP consultations before 2013 are greyed-out to indicate that they occurred before PrEP guidelines came into effect (these were also excluded from subsequent analyses).

From January 2013 onwards, of the 2,682 participants who received nPEP and who met the inclusion criteria (i.e., gbMSM, cis-male, consented participation), 36% (972/2,679) also consulted for or received PrEP during that period and only 1% (13/972) reported having had a PrEP consultation outside of l’Actuel. Nearly half of the 2,718 consenting gbMSM PrEP participants also used nPEP during that period (46%, 1,242/2,715), 10% of whom (120/1,242) received nPEP externally to l’Actuel. The majority of participants in both the nPEP and PrEP cohorts consulted for nPEP prior to PrEP (90%, 861/959).

Almost all *PEP-to-PrEP* participants were prescribed PrEP (98%, 847/861) and 17% (144/861) reinitiated nPEP afterwards (Figure 1).

#### Participants included in the main analysis

Overall, 1,707 *PEP only* participants, 861 *PEP-to-PrEP* participants and 1,473 *PrEP only* participants were included in the main analyses. More *PEP-to-PrEP* participants had a history of multiple nPEP episodes than *PEP only* participants. Furthermore, *PEP-to-PrEP* participants reported a 6-month shorter average time between nPEP episodes than *PEP only* participants. Approximately 44% (382/861) and 24% (414/1,707) of *PEP-to-PrEP* participants and *PEP only* participants had at least 2 nPEP episodes.

Additionally, the proportion of participants returning for the first nPEP follow-up visit and who were adherent to nPEP treatment was 19% and 12% higher among *PEP-to-PrEP* participants than *PEP only* participants. Compared to the other 2 groups, *PrEP only* participants had a lower education level and the proportion self-reporting a lifetime history of STIs was 10% higher (**Table 1**).

**Table 1.**
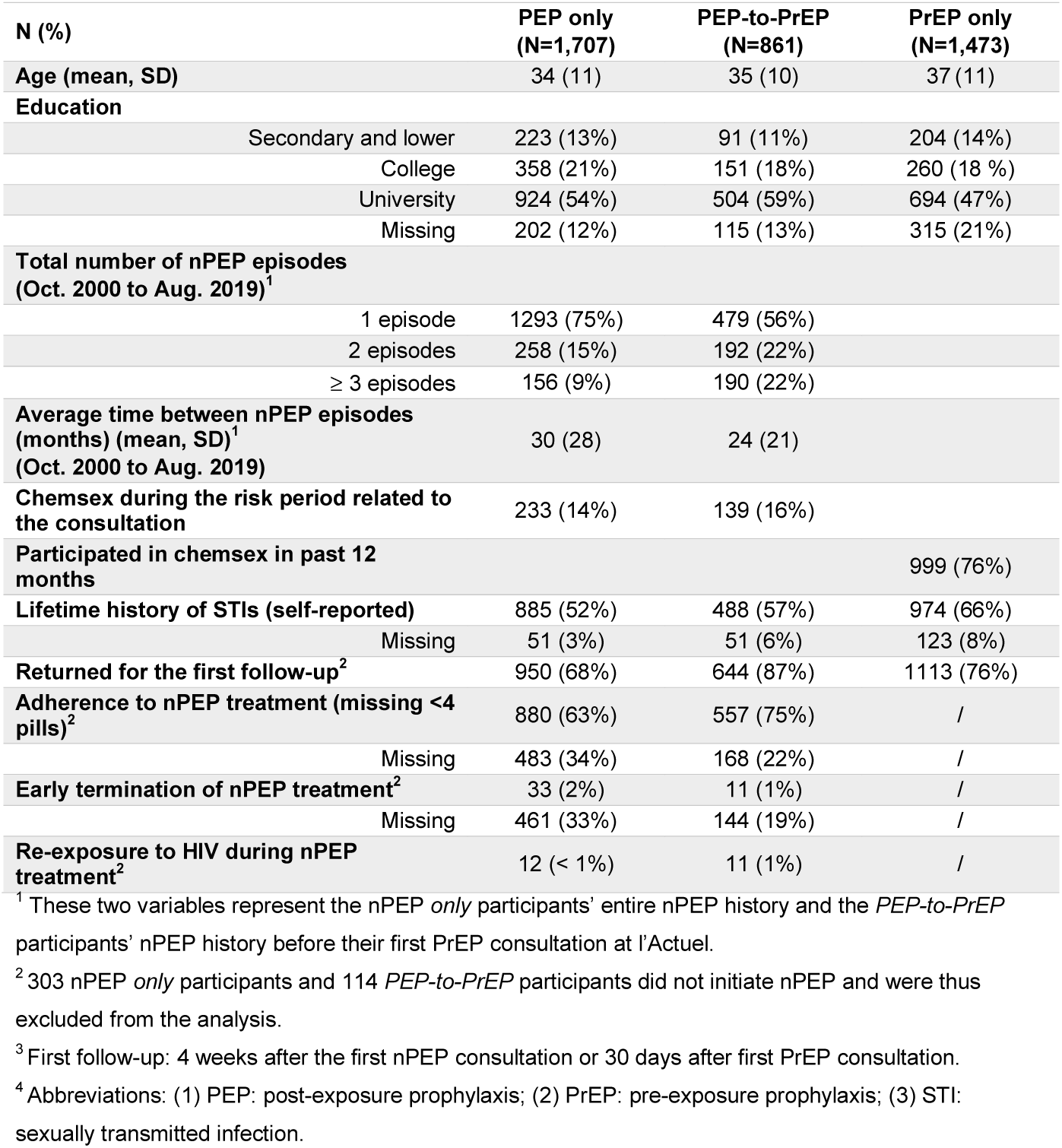
Demographic and sexual behaviour characteristics of gay, bisexual, and other men who have sex with men (gbMSM) attending *Clinique médicale l’Actuel* at first nPEP consultation or PrEP consultation after 2013 for nPEP *only, PEP-to-PrEP*, and *PrEP only* participants.

### Time from nPEP consultation to PrEP uptake and its determinants

During the study period, approximately 40% of the nPEP participants linked to PrEP. One quarter of the participants linked to PrEP within the first 20 months after first nPEP consultation. Participants younger than 25 years old had lower rates of PrEP linkage years and participants who had at least 2 nPEP episodes had overall higher rates of PrEP linkage. Participants with more nPEP episodes linked to PrEP at a higher rate. The rate of PrEP linkage increased over time between 2013 and 2019 (**Figure 3 A, B, C**).

**Figure 3.**
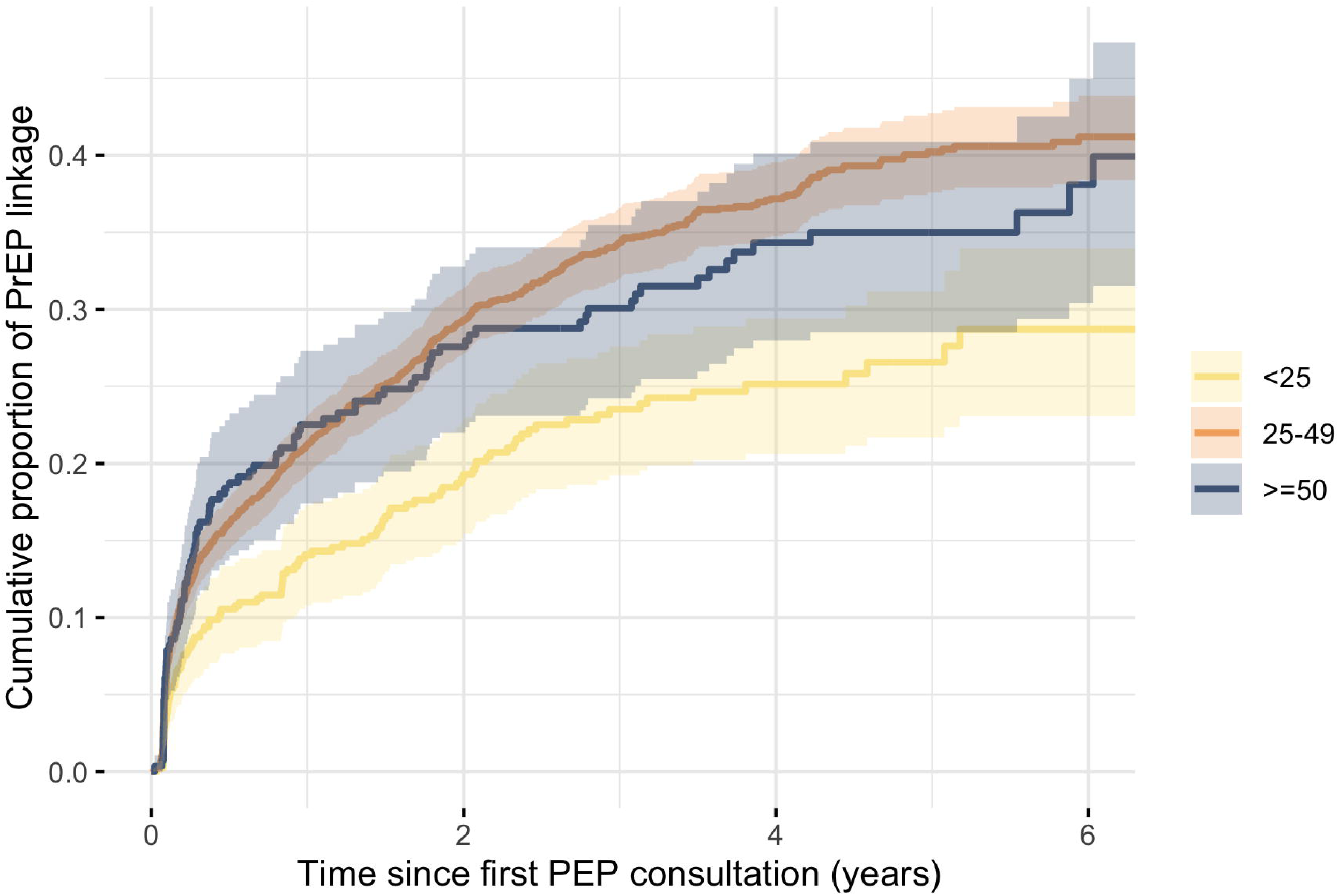
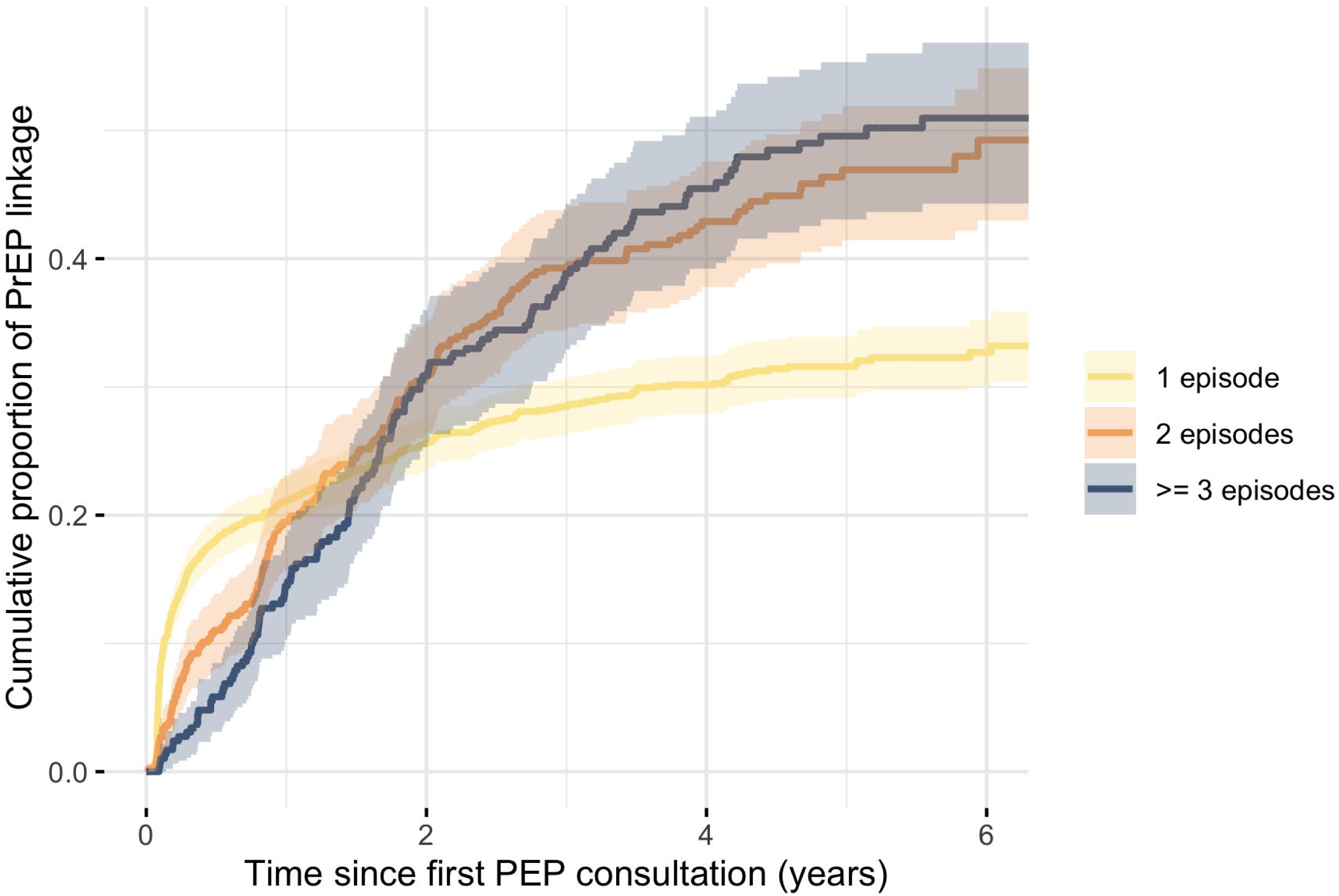
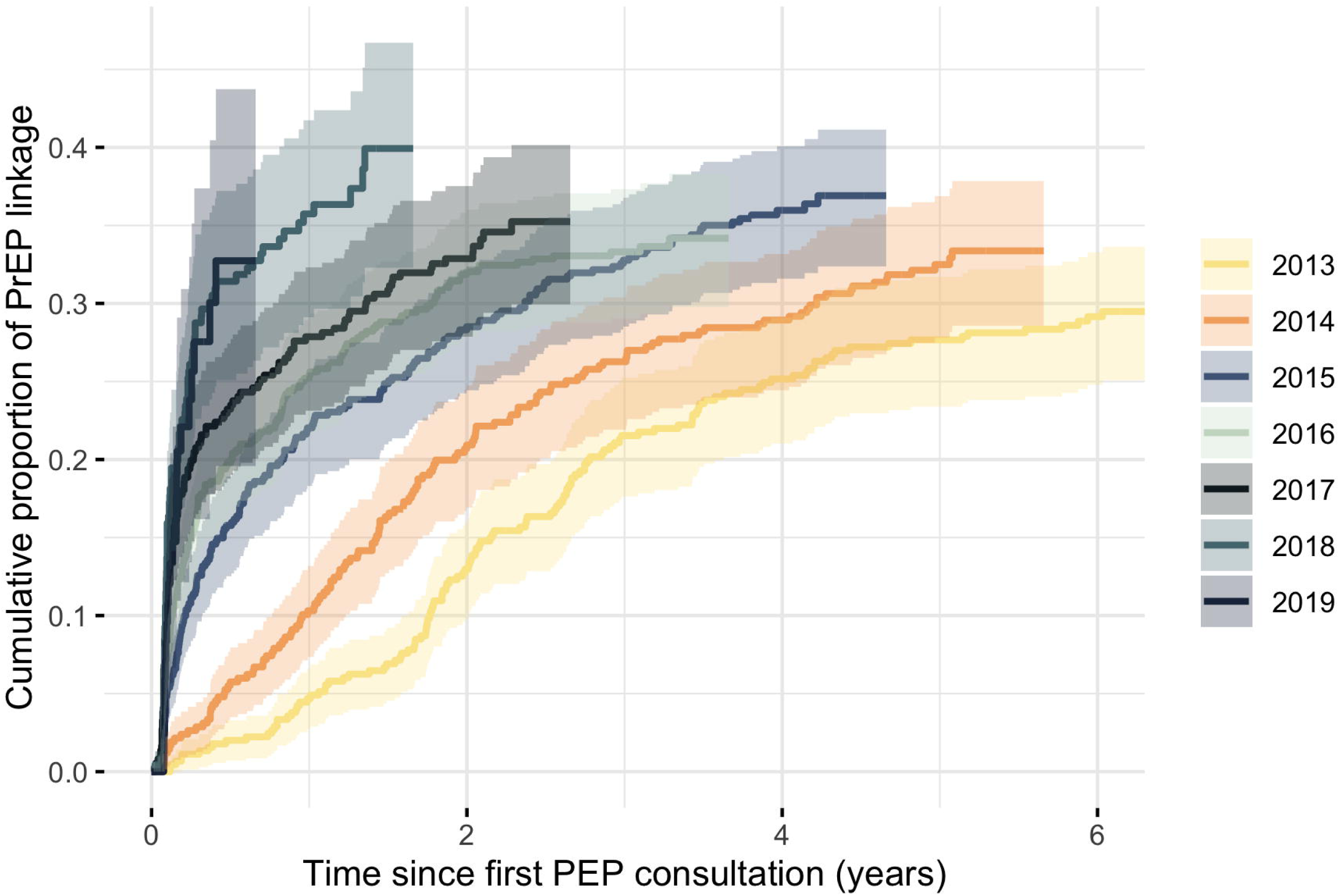
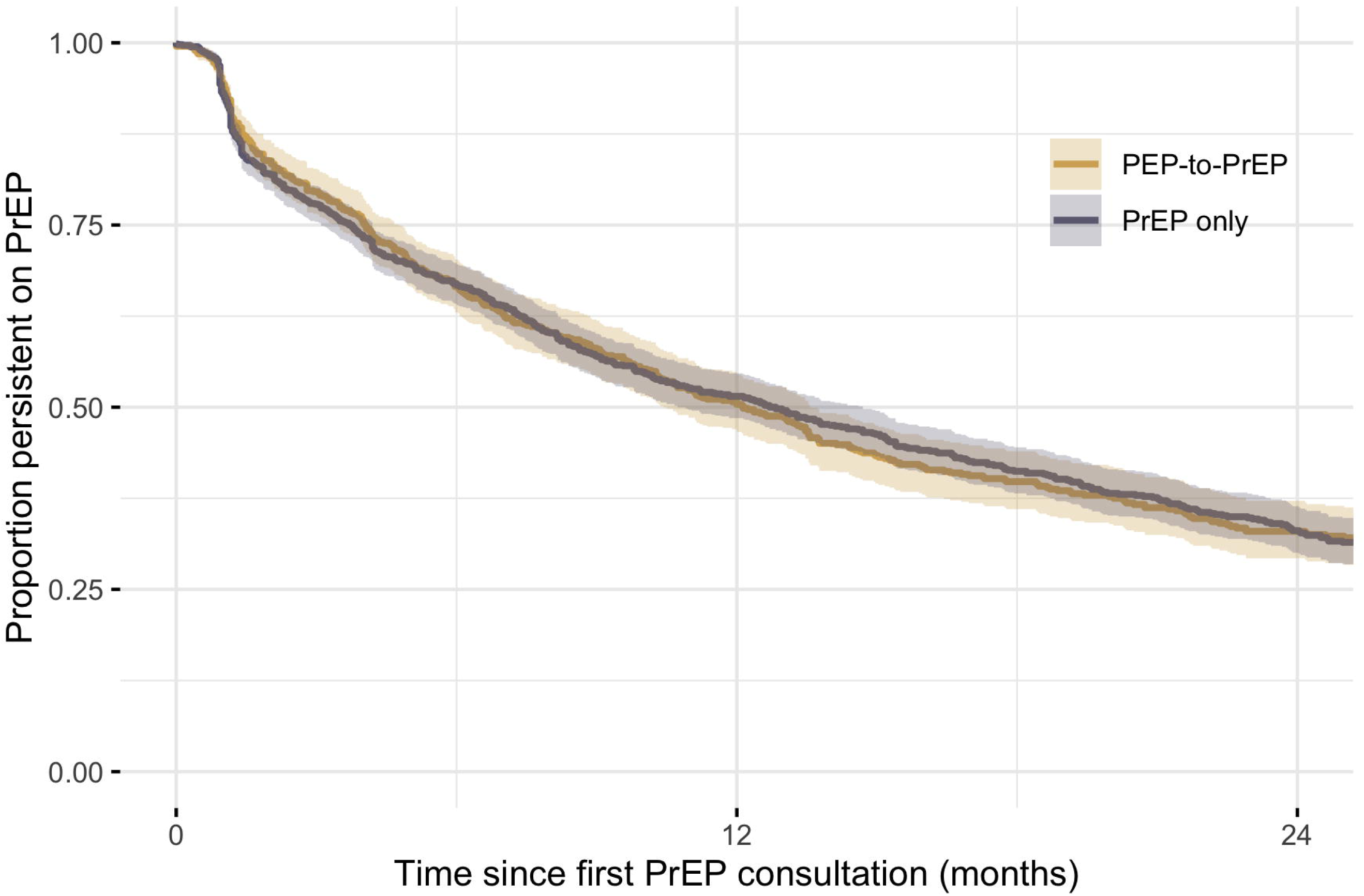
Survival curves of: (1) The cumulative proportion and 95% confidence intervals for consulting for pre-exposure prophylaxis (PrEP) after a post-exposure prophylaxis (PEP) consultation stratified by age group (<25, 25-49, and ≥ 50 years) (Panel A), stratified by number of nPEP episodes (1 episode, 2 episodes, and ≥3 episodes) (Panel B), and stratified by calendar year of first nPEP consultation (Panel C), and (2) the proportion of participants who were persistent with pre-exposure prophylaxis (PrEP) stratified by post-exposure prophylaxis (PEP) history (Panel D) at Clinique médicale l’Actuel among gay, bisexual and other men who have sex with men (gbMSM). The x-axis represents the time since first nPEP consultation (Panels A, B, C) or time since first PrEP consultation (Panel D) after 2013 at *Clinique médicale l’Actuel*. PEP-to-PrEP: participants who previously consulted for nPEP and initiated PrEP at *Clinique médicale l’Actuel*. PrEP only: participants who self-reported never having consulted for nPEP (anywhere) but initiated PrEP at *Clinique médicale l’Actuel*.

Participants aged 25-49 years of age, who had multiple nPEP episodes, chemsex during the risk period, self-reported antecedent STIs, and returned for the first nPEP follow-up visit were the main determinants of time to PrEP linkage in the multivariate model (**Table 2**). The association between education and PrEP linkage was inconclusive although participants with a university degree had a slightly higher hazard ratio than those who finished college. Estimates of the total number of nPEP episodes suggested that each additional nPEP consultation would increase the rate of PrEP linkage among nPEP users by 39% (HR=1.39, CI: [1.31-1.46]). Chemsex during the risk period and having antecedent STIs increased the rate of PrEP linkage by 30% (HR=1.30, 95% CI: [1.08, 1.56]) and 46% (HR=1.46, 95% CI: [1.26, 1.69]), respectively. Returning for the first nPEP follow-up visit at week 4 was strongly associated with linkage to PrEP, increasing the rate of linkage to 3.36 (95% CI: [2.71, 4.16]) times the rate for those who did not return for the follow-up visit.

**Table 2.** Univariable and multivariable hazard ratios and 95% confidence interval (95%CI) from Cox proportional hazard models of the determinants of time to PrEP consultation among gay, bisexual, and other men who have sex with men (gbMSM) who used nPEP after January 2013 at the *Clinique médicale l’Actuel*.

| Determinants | Univariate analysis<br>(N=2,571) |  | Multivariate analysis<br>(N=2,571) |  |
| --- | --- | --- | --- | --- |
|  | Hazard ratio | 95%CI | Hazard ratio | 95%CI |
| **Age (years)** |  |  |  |  |
| < 25 | REF |  | REF |  |
| 25 - 49 | 1.61 | [1.31, 1.98] | 1.33 | [1.08, 1.65] |
| ≥ 50 | 1.47 | [1.11, 1.95] | 1.08 | [0.81, 1.44] |
| **Education<sup>1</sup>** |  |  |  |  |
| Secondary | REF |  | REF |  |
| College | 1.01 | [0.77, 1.31] | 0.96 | [0.73, 1.27] |
| University | 1.24 | [0.98, 1.56] | 1.04 | [0.82, 1.32] |
| **Total number of nPEP episodes** | 1.34 | [1.25, 1.44] | 1.39 | [1.31, 1.46] |
| **Chemsex during the risk period related to PEP consultations** | 1.23 | [1.02, 1.49] | 1.30 | [1.08, 1.56] |
| **Self-reported lifetime STI history<sup>1</sup>** | 1.54 | [1.34, 1.77] | 1.46 | [1.26, 1.69] |
| **Return for the first nPEP follow-up visit<sup>1</sup>** | 3.09 | [2.49, 3.83] | 3.36 | [2.71, 4.16] |

<sup>1</sup> Due to missing values, education, self-reported lifetime STI history and return for the first follow-up visit were imputed using 5 multiple imputations. Pooled estimates based on Rubin's rules are shown in the table.
<sup>2</sup> Abbreviations: (1) PEP: post-exposure prophylaxis; (2) STI: sexually transmitted infection.

### PrEP persistence among PEP-to-PrEP and PrEP only participants

Daily PrEP was prescribed to 78% (674/861) of the *PEP-to-PrEP* participants and 82% (1,208/1,473) of the *PrEP only* participants while the remainder of the participants were prescribed intermittent PrEP. Following their PrEP consult, 78% of the *PEP-to-PrEP* participants and 76% of the *PrEP only* participants initiated PrEP. The probability of PrEP persistence was similar between *PEP-to-PrEP* and *PrEP only* participants (Figure 3D). The median time to PrEP discontinuation was 0.59 (95% CI: [0.06, 3.44]) and 0.54 years (95%CI: [0.06, 2.96]) among *PEP-to-PrEP* and *PrEP only* participants, respectively. Half of the participants discontinued PrEP within the first year of initiation.

## Discussion

PrEP is a highly effective biomedical HIV prevention method recommended for high-risk populations, such as gbMSM, and is an important component of combination prevention strategies to achieve HIV elimination [4 6 12]. In our analysis of six years of longitudinal data from the l’Actuel nPEP and PrEP cohorts, approximately 40% of nPEP users were linked to PrEP, of which one quarter of the linkages occurred within the first 20 months after nPEP consultation. nPEP participants aged 25-49 years, who reported multiple nPEP episodes, chemsex, antecedent STIs, and who returned for the first nPEP follow-up visit were more likely to link to PrEP. Reporting a history of nPEP use did not impact PrEP persistence.

Our evaluation suggests that the time from nPEP consult to PrEP linkage rapidly improved over the 2013-2019 period. This was particularly evident after 2015 when more than 20% of individuals consulted for PrEP within the first year of their nPEP consult. A study conducted in Toronto by Siemieniuk *et al*. (2005) indicated that 10% of the individuals who consulted for nPEP at the *HIV Prevention Clinic* between 2013 and 2014 initiated PrEP as of 2015 [24]. This proportion is much lower than what was found here-but could be explained by the lack of a publicly funded PrEP program in Ontario at the time.

Despite improvements in PEP-to-PrEP linkage at l’Actuel, such progress could be compromised by the overall low persistence on PrEP. Indeed, half of the PrEP users discontinued it during the first year. There are multiple reasons for the observed PrEP discontinuation including: changes in sexual behaviors, such as increased condom use, and the formation of a single long-term partnership [25 26]. However, the discontinuations could be concerning if they are not related to change in sexual behaviors. There is some evidence to suggest that this is not always the case. For example, 98 (7%) PrEP users, and 144 (17%) *PEP-to-PrEP* users sought nPEP after PrEP discontinuation. If this group is composed of individuals with a low-frequency of high-risk exposures, other interventions such as PIP (post-exposure prophylaxis in pocket) should be considered [27]. This could be important for PEP-experienced users. A recent study in Toronto recorded a high prevalence of syndemic health problems among MSM seeking nPEP [28]. This clustering of syndemic conditions could partly explain low persistence on PrEP of nPEP users, which was also observed in other cohorts [29].

Our results should be interpreted considering several limitations. First, we cannot rule out that participants who used nPEP at l’Actuel eventually consulted for PrEP at other clinics. As such, we may have overestimated time to linkage. Despite this limitation, only 1% of participants in the nPEP cohort reported having taken PrEP outside of l’Actuel. Further, only 10% of the PrEP cohort had consulted for nPEP at another clinic. This could imply that our inability to link our nPEP and PrEP cohorts to other clinics may have only small impacts our inferences. However, given the changing HIV prevention landscape in Montreal after 2017, future work should include multi-sites collaboration. Second, it is difficult to know the exact date of PrEP termination for participants who did not return for a follow-up visit which may result in incorrectly estimating the participants’ adherence to a PrEP regimen. The date of last PrEP consultation was used as the end date of PrEP for those who discontinued PrEP, which might underestimate the time on PrEP. Additionally, participants on an intermittent PrEP regime may have sufficient pills to last for more than 6 months, prolonging the time between follow-up visits, which would be treated as “terminated PrEP” in this study and lower the estimated PrEP persistence. However, as the PrEP persistence analysis performed in this study was aimed to observe the difference between PrEP users with and without a nPEP history, this could have minimal impact on our results. Third, and not unlike other studies, we relied on self-reported information regarding potentially sensitive behaviors (e.g., antecedent STI, sexual behaviors, sexualized substance use) that could have been misreported. Finally, the number of missing values for certain covariates was non-negligible. However, multiple imputation was used which efficiently propagated uncertainty to the relevant effect size measures.

As one of the largest nPEP and PrEP providers in Canada, the data collected at *Clinique médicale l’Actuel* provides a comprehensive data set characterising the trends in nPEP and PrEP use among gbMSM in Montréal. The large sample size of this study enabled us to conduct subgroup analyses. Additionally, the study period covered the time from the beginning of PrEP implementation in Québec until the present which made it possible to capture a complete picture of PrEP use development and the impact of PrEP on nPEP use. These strengths have resulted in a robust quantification of the relationship between nPEP and PrEP use among gbMSM in this Canadian setting.

## Conclusions

Making PrEP accessible, and promoting its use, to high risk populations is essential to prevent HIV transmission. Understanding the current gaps and the determinants of PEP-to-PrEP linkages could help optimize PrEP delivery. Our results suggest that creating a strong linkage corridor between nPEP and PrEP services could help facilitate PrEP uptake among gbMSM vulnerable to HIV acquisition. Furthermore, given that 17% *PEP-to-PrEP* users sought nPEP after PrEP discontinuation, future interventions should focus on increasing PrEP continuation among people at on-going HIV risk. These results should be used to better inform individualized HIV prevention strategies if Montréal’s Fast-Track city goal of HIV elimination is to be achieved, contributing to the global effort to eliminate HIV/AIDS by 2030 [30].

## Data Availability

The data that support the findings of this study are available from Clinique Medicale l'Actuel but restrictions apply to the availability of these data, which were used under license for the current study, and so are not publicly available.

